# Prognostic Markers of Frailty and Fall Risks in Older Adults: a Longitudinal Cohort Study using Explainable AI

**DOI:** 10.64898/2026.03.13.26347338

**Authors:** Tiago Nóbrega, Tiago Santos, Henrique Anjos, Bruno Gomes, Fábio Cunha, Pedro Oliveira, Rui Baptista, Andreia Pizarro, Jorge Mota, Daniel Moreira-Gonçalves, Rui Henriques, Rafael S. Costa

**Affiliations:** UCIBIO & i4HB, NOVA School of Science and Technology, NOVA University, Portugal; INESC-ID and Instituto Superior Técnico, Universidade de Lisboa, Portugal; Municipality of Vila Nova de Famalicão, V. N. Famalicão, Portugal; CIAFEL, Faculty of Sport, University of Porto and ITR, Porto, Portugal

**Keywords:** Frailty, Fall Risk, Geriatric Health, Handgrip Strength, Explainable AI, Prognosis, Risk Stratification

## Abstract

Frailty is a geriatric syndrome that reflects a state of increased vulnerability to endogenous and exogenous stressors, exposing individuals to a higher risk of premature death and adverse health outcomes. This study aims at identifying novel determinants of frailty and refining screening tools to improve the prognostication of understudied endpoints. To this end, we established a new longitudinal cohort study comprising 2,862 participants and 6,855 observations across up to four assessment moments, incorporating measures of anthropometry, functional performance, cognition, quality of life, fall history, and sociodemography. We combine unsupervised clustering to explore profile heterogeneity with supervised prediction for falls, hospitalization, and handgrip strength, using explainability machine learning approaches to connect model outputs to clinically meaningful patterns. Outcome-agnostic clustering separates functional profiles that align with different fall burdens, while a complementary outcome-aware analysis provides an integrated stratification combining functional limitations and perceived risks. Predictive models achieve moderate and consistent discrimination for fall prediction (AUROC = 0.636–0.696), and explainability approaches consistently emphasize key drivers including handgrip strength, self-report assessments, and other results from functional tests. Handgrip regression attains MAE ≈ 3.6 kg (*R*^2^ ≈ 0.49), while a dedicated CatBoost sarcopenia classifier improves detection (AU-ROC = 0.798, recall = 0.792) at the cost of low precision, consistent with screening-oriented use. Overall, the results support the feasibility of explainable AI for actionable risk stratification in community assessments, while highlighting constraints related to missingness, class imbalance, and healthy volunteer bias.

## 1 Introduction

Population ageing is reshaping health-care systems worldwide, and frailty is increasingly recognised as an important public health concern with major implications for clinical practice and health systems [20]. In Portugal, this challenge is particularly salient, where an estimated 15% of older adults live with the syndrome [29], one of the highest percentages in the EU and escalates further with advancing age [16]. Moreover, many older cohorts have low formal education, which can compound social and health vulnerability and influence access to prevention and care [15]. Given the increasing burden of age-related vulnerability, effective stratification and prognosis of frailty risks are aligned with key primary healthcare reforms designed to strengthen preventive care (e.g., [13]).

Frailty is a multidimensional geriatric syndrome distinct from chronological ageing, multimorbidity, and disability, although it frequently coexists with these conditions. It is characterised by diminished physiological reserve and impaired homeostatic resilience across multiple organ systems, resulting in increased vulnerability to external stressors and adverse outcomes [24, 40]. As a dynamic condition that exists along a continuum from robustness to severe frailty, frailty is driven by complex interactions among biological (e.g., sarcopenia, polypharmacy risks, dysregulation of inflammatory, endocrine and immune pathways), behavioural (e.g., physical activity, nutritional intake) and psychosocial determinants [3, 24]. Transitions between states are possible, particularly at earlier stages (pre-frailty), and multicomponent interventions, especially those combining resistance exercise, nutritional optimisation, and comprehensive medication review, have demonstrated potential to stabilise or improve frailty status, highlighting the role of tailored preventive practices [24].

Two complementary conceptual models of frailty predominate in the literature: the physical phenotype model, which operationalises frailty through observable features such as weakness, slowness, exhaustion, weight loss, and low physical activity; and the deficit accumulation model, which conceptualises frailty as the cumulative burden of health deficits across clinical, functional, and psychosocial domains [24]. Both frameworks capture higher susceptibility to falls and fractures, hospitalisation, reduced quality of life, institutionalisation, and premature mortality [24]. To operationalize frailty risk stratification in routine care, classical scoring systems derived from routinely collected records have been proposed, including the electronic Frailty Index (eFI) for primary care and the Hospital Frailty Risk Score (HFRS) for secondary care settings [8, 18]. However, as these tools are largely driven by coded deficits and administrative diagnoses, they generally neglect individual-level characteristics (e.g., functional performance) needed for actionable screening and prognostic decisions; consequently, they are best viewed as complementary population-scale screening instruments rather than standalone, individualized clinical decision support.

In this context, machine learning offers a way to integratively model heterogeneous clinical, functional and sociodemographic data to support earlier identification and stratification of individuals at risk. Despite the growing studies on machine learning for frailty assessment, several important challenges remain. Many existing studies rely on relatively small or cross-sectional cohorts, generally neglecting the multifactorial nature of frailty, focusing on a single clinical endpoint, or treating frailty as a predefined diagnostic label, overlooking its heterogeneous and dynamic components. Furthermore, predictive performance is frequently prioritised over clinical interpretability, limiting the translation of these methods into routine healthcare practice. In this context, there is a need for approaches that simultaneously tap into multimodal and longitudinal stances for the identification of actionable risk profiles beyond predefined diagnostic categories, prognosis of clinical endpoints of interest, and transparent explanations of the factors driving model predictions.

To address these challenges, this study introduces a large-scale cohort study conducted by the Municipal Council of Vila Nova de Famalicão that is uniquely characterized by a broad, multimodal follow-up battery of clinical, functional, and self-reported assessments, collected repeatedly across several time points for 2,862 older adults, enabling both cross-sectional profiling and longitudinal screening of functional change. Using this cohort study, this work proposes an explainable machine-learning framework for an actionable stratification of frailty-related vulnerability profiles and prognostication of clinically relevant outcomes, including fall-related risks and sarcopenia.

The main contributions of this work are twofold: i) a predictive study addressing the prognosis of multiple clinically relevant endpoints, with a particular emphasis on model explainability to facilitate knowledge acquisition and support clinically grounded interpretation of new putative determinants; ii) a descriptive study to explore frailty heterogeneity and identify actionable profiles under both cross-sectional and longitudinal stances, with the primary goal of linking model outputs to clinically interpretable patterns.

## 2 Related Work

Frailty, being a syndrome rather than a disease, remains difficult to define with close consensus, since it spans multiple domains and manifests through heterogeneous deficits across individuals [21]. This conceptual heterogeneity is reflected in practice by the coexistence of multiple frailty measurement approaches with different assumptions, inputs, and intended uses [14]. Moreover, although frailty is broadly understood as age-associated vulnerability, a key challenge lies in how to operationalise it consistently in clinical practice and research, which has motivated deficit-accumulation formulations such as the Frailty Index [32]. Consequently, developing machine-learning models for cross-study analysis is more challenging since the target clinical endpoints and the input feature spaces largely depend on the chosen operational definition and measurement pipeline, which can limit comparability and generalisability.

Recent work has applied machine learning across multiple, complementary targets relevant to geriatric vulnerability. For fall-risk prediction, short-term models based on routinely collected electronic health records leverage dynamic physiological signals (e.g., blood pressure and other vital signs) to improve surveillance in long-term care settings [36]. Interpretability has also been used to compare how the predictive feature landscape changes across clinically meaningful subgroups. For example, separate fall-risk models for people with versus without pain highlight pain characteristics and laboratory blood markers as relevant predictors in the pain group, suggesting that pain not only increases risk, but can also shift which factors matter the most for prediction [5]. In parallel, frailty prediction has been explored using machine-learning models for multiple adverse frailty-related outcomes(e.g., mortality, hospitalizations), supporting the development of decision-support tools for early identification [35]. Complementary studies emphasize multidomain inputs and explainable AI in community settings, combining clinical, functional, lifestyle, social, and digital indicators with feature-attribution methods to support actionable screening [25]. Functional markers such as handgrip strength are widely used as practical indicators of age-related physical decline and are central to sarcopenia and frailty assessment. Accordingly, recent work has modelled grip-strength within interpretable pipelines, while complementary studies frame sarcopenia prediction using explainable models that incorporate clinical and blood-based variables to contextualize muscle-strength decline [7, 41]. Overall, across the AI literature on frailty identification and diagnosis, studies target a wide range of labels and outcomes and rely on heterogeneous feature sources (e.g., clinical records, functional tests, sensors, and sociodemographic variables), yet a consistent theme is the emphasis on explainability across all studies [38].

Given the absence of an explicit frailty diagnosis variable in our dataset, the aim of this work is to assess the predictive value of the monitored features considering their high-order dependencies, produce actionable explanations, and identify what can be inferred from the available longitudinal assessments. By avoiding reliance on a single frailty definition, we reduce the risk of bias tied to any specific diagnostic framework and avoid over-reliance on a single scale. However, we acknowledge that this choice introduces additional uncertainty, since no single ground-truth frailty proxies, such as handgrip strength tests, are selected. Within this framing, we combine unsupervised clustering to study heterogeneity and uncover subgroup profiles with supervised prediction tasks focused on actionable endpoints, namely fall-risk prediction and sarcopenia-related risk, while using explainability to connect model outputs to clinically meaningful patterns.

## 3 Methods

### 3.1 Dataset and Outcomes

The undertaken study employed a combined retrospective–prospective cohort design within Famalicão’s municipal program *Mais e Melhores Anos* (https://www.famalicao.pt/mais-e-melhores-anos) from November 2021 to June 2023, where older adults underwent a comprehensive battery of health tests, and the data were subsequently analyzed retrospectively. The aim of the municipal program is to promote and evaluate the impact of regular physical activity on the health of the elders, with participants being periodically reassessed in a biyearly modality [1]. The anonymized dataset was provided by the Municipality of Vila Nova de Famalicão.

In each assessment time point, older adults are characterized through the following domains of variables: i) Sociodemographic and program-related: age, sex, education, attendance moment; ii) Anthropometry: height, weight, body composition; iii) Physical activity: type of sport practiced and frequency; iv) Functional tests: handgrip strength, Timed Up and Go (TUG), sit-to-stand, 6-minute walk; v) Cognitive: Mini-Mental State Examination (MMSE); vi) Quality of life: EuroQol 5-Dimension 5-Level (EQ-5D-5L) and vii) Fall history: How many falls, in what circunstances, and their consequences.

Besides the quantitative test outcomes, the protocol also records complementary contextual information as non-structured fields, such as the circumstances of a fall or the reason why a participant stopped during the 6-minute walk test.

Falls were considered the main outcomes in this study, focusing on both their occurrence over the next 6 months (next time point assessment) and severity-related consequences, namely hospital assistance and hospitalizations. Since the latter two outcomes are comparatively rare and therefore more challenging to model, they were treated as secondary outcomes. In addition, handgrip strength—or probable sarcopenia derived from handgrip strength, by applying the EWGSOP2 cut-off—was treated as a secondary outcome in a separate task from the remaining assessment variables.

All the code developed in this study for modeling and analysis is freely available at https://github.com/tiagonob80/FRAIL-public and the dataset is available upon reasonable request.

### 3.2 Exploratory Data Analysis

A total of 2,862 participants were included in the study, yielding 6,855 observations, with 767 participants contributing with a single recording session, 740 with two records, 812 with three records, and 543 with all four records conducted along the 2-year span of the study. The mean participant age is 69.89 years, and the sample includes 68% women and 32% men.

Participation records indicate a highly active cohort: 94% of participants practice physical exercise twice per week and 92% report swimming activities. Given that the 1h/session municipal program explicitly promotes regular physical activity, the sample can be considered predominantly active, which is also reflected in fall circumstances frequently involving demanding activities (e.g., running or climbing ladders) that are more typical of healthier and functioning older adults.

In this sample, 36% of people report having fallen at least once, with 11% requiring hospital assistance and 2% reporting hospitalization stay due to a fall.

Older people can also be characterized by the EQ-5D-5L questionnaire, which captures health-related quality of life across five dimensions (mobility, self-care, usual activities, pain/discomfort, and anxiety/depression). These are rated 1–5, where 1 indicates no problems and higher levels indicate increasing severity of problems. The questionnaire also includes a self-rated health visual analogue scale (0—100).

In the mobility domain, 85% of respondents reported little-to-no problems walking (levels 1–2). In the self-care domain, 96% reported little-to-no problems with dressing or bathing. Similarly, 94% reported little-to-no limitations in usual activities, whereas pain/discomfort and anxiety/depression showed comparatively higher impairment, with 83% and 88% reporting levels 1–2, respectively.

The visual analogue scale (Fig.2) portrays a predominantly active and functioning older cohort, with generally favorable self-reported quality of life despite a non-negligible burden of pain/discomfort and a substantial prevalence of falls (in the first assessment moment, 22.4% of the 1417 participants reported at least one fall in the past 12 months).

**Figure 1.**
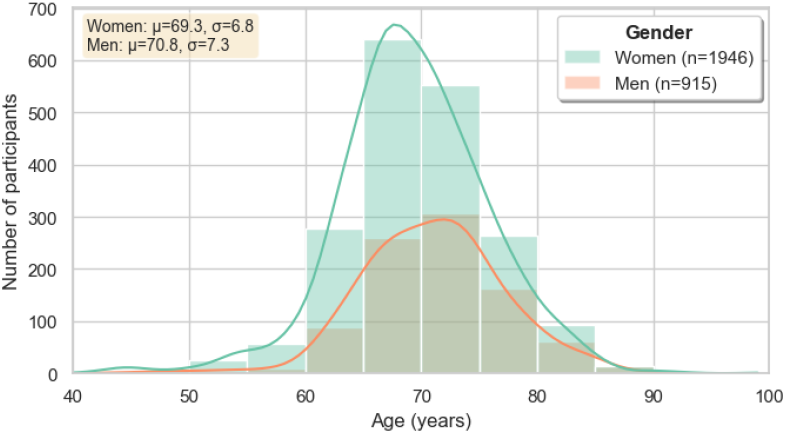
Cohort Statistics: age distribution by gender.

**Figure 2.**
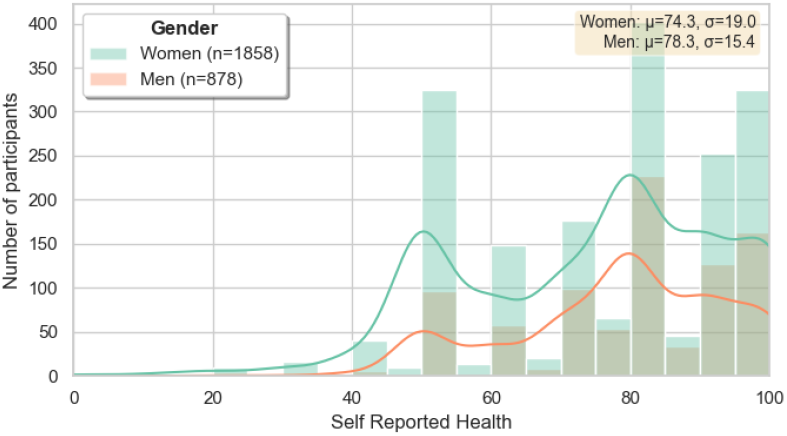
Self Reported Health (0–100) per participant using the most recent available response.

While the previous exploratory characterization is based on the available (non-missing) values, the collected data contains non-uniformly distributed missing values across variable groups. The sociodemographic and anthropometric fields are largely complete, except the visceral fat measure, which has approximately 24% missing entries. The outcome variables, namely fall history, their consequences and handgrip strength, are also complete.

Some variable groups show higher levels of missingness. The EQ-5D-5L dimensions present around 20% missing values, and the MMSE items exhibit a similar missingness rate.

#### 3.2.1 Normative Benchmarking and Cohort Selection Bias

Since the dataset was collected within a municipal physical-activity context, it likely over-represents older adults who are healthier and more active than the general population, which can induce selection bias and systematic deviations from population norms. The objective of this normative benchmarking is therefore to quantify such potential bias by measuring how far the cohort departs from Portuguese reference values across key physical-performance and anthropometric indicators.

For each outcome, participants values are matched to the appropriate normative stratum and compared to Portuguese reference values: handgrip strength is benchmarked using sex- and age-specific values stratified by sex-specific height tertiles [30], while functional-fitness tests (chair stand repetitions, 6-min walk distance, and 8-ft up-and-go time) were benchmarked using sex- and age-interval norms (60–64, 65–69, 70–74, 75–79, +80) [19]. Stratum-specific z-scores are computed and tested on their central tendency differed from zero (one-sample t-test for mean z=0 and Wilcoxon signed-rank for median z=0). Overall, the cohort exhibited better performance in strength/endurance-oriented tests (chair stand and 6-min walk) and a modest upward shift in handgrip, consistent with a healthier/more active sample and therefore a plausible selection bias. Conversely, up-and-go times appeared higher than the reference, however, this result should be interpreted cautiously because time-based mobility tests are highly sensitive to protocol configuration (e.g., course length, chair height, turning rules, timing method, and allowance of aids), and the local test setup presented may not be directly comparable to the reference protocol [19].

Taken together, these comparisons suggest that the participants constitute a generally healthier and more physically capable cohort than the normative samples, which supports the presence of selection bias toward fitter individuals. This bias should be explicitly acknowledged when interpreting downstream analyses and when generalizing findings to the broader older-adult population.

### 3.3 Preprocessing

#### 3.3.1 Text standardization

Several of the monitored features were manually entered during each live assessment, leading to heterogeneity in some of the binary, categorical and free-text response fields. To reduce artificial variability (e.g., spelling inconsistencies, capitalization, and abbreviations) and ensure consistent encoding across moments and participants, a text standardization step was applied to focus on non-numeric fields. Semantically equivalent responses were harmonized, such as mapping multiple forms of affirmative/negative answers (e.g., “S”, “Sim”, “SIM” vs. “N”, “Não”, “NÃO”) into a single representation (“sim”/”não”). Additionally, missing information appeared under diverse representations (e.g., empty strings, “N/D”, “N/A”, “ND”, “NA”) depending on the assessor. All such cases were consistently converted to NaN, ensuring that “unknown” values are standardly represented and can be handled downstream in a principled manner.

Finally, some variables contained short unstructured descriptions (e.g., fall circumstances) whose raw motifs exhibited high cardinality. For these fields, we performed a lightweight normalization based on keywords to consolidate frequent patterns into a smaller set of interpretable categories (e.g., mapping mentions of stairs, ladders, walking, running, slipping, or imbalance to standardized labels). Overall, this step reduced noise and cardinality in textual features while preserving their semantic content, resulting in more consistent inputs for subsequent analysis and modeling.

#### 3.3.2 Outlier detection and treatment

This step entailed correcting or removing anomalous values caused by measurement/recording errors, transcription mistakes, and inconsistencies across assessment moments. Through the longitudinal nature of the data, repeated measurements across moments can be used to detect implausible shifts in age, height and weight. Invalid values were corrected using the participant’s own valid records; otherwise, they were set to missing (NaN) or the corresponding subject was excluded from the analysis.

For **age**, we firstly considered only as valid entries people with 40–110 years of age. Since the 4 moments of assessment occurred within 2 years, participants with excessive age variation were removed, as this strongly indicates ID linkage or data-entry issues. For **height** and **weight**, unusually large within-subject shifts were flagged and, when there were at least three valid measurements for that participant, the single most discrepant observation (largest absolute deviation from the participant median) was set to NaN, to later be filled.

Beyond removing erroneous entries, the impact of extreme values was reduced via capping/clipping using heuristic limits informed by variable distributions and domain knowledge. A Tukey rule (e.g., *Q*_1_ − 3 · *IQR* / *Q*_3_ + 3 · *IQR*) was tested but proved too aggressive, collapsing variability and producing many identical capped values. Instead, we applied smoother, Winsorization-style bounds tailored per variable group, which reduces the influence of outliers while preserving most of the original information. This method generally improves K-means clustering stability [23]. In this context, anthropometry measures were bounded to reduce extreme values; handgrip attempts were capped at 60kg; 6-minute walk derived measures were capped (e.g., number of laps, total distance); MMSE sub-scores were clipped to their valid score ranges, as well as EQ-5D-5L dimensions.

This pipeline produced a more reliable dataset, correcting inconsistent measurements, setting irrecoverable values to missing, and excluding a small number of participants with patterns suggestive of entry errors. The domain-informed clipping reduced the leverage of extreme values, aiding the stability of the prospective distance-based modeling and clustering, while preserving most of the original variability compared with the overly aggressive IQR-based rule.

#### 3.3.3 Missing data analysis and treatment

An imputation pipeline combining longitudinal within-subject filling, rule-based completion for logically dependent fields, and multivariate imputation is undertaken to reduce missingness while preserving internal consistency for downstream analyses.

A large share of missings are meaningful and structurally driven: several fields are only applicable when the corresponding event occurs (e.g., stopping during the 6-minute walk test or hospitalization related variables). Therefore, high missing rates in these variables predominantly reflect participants who did not experience the event, rather than data-collection failures. Figure 3 shows the percentage of missing values per variable section.

**Figure 3.**
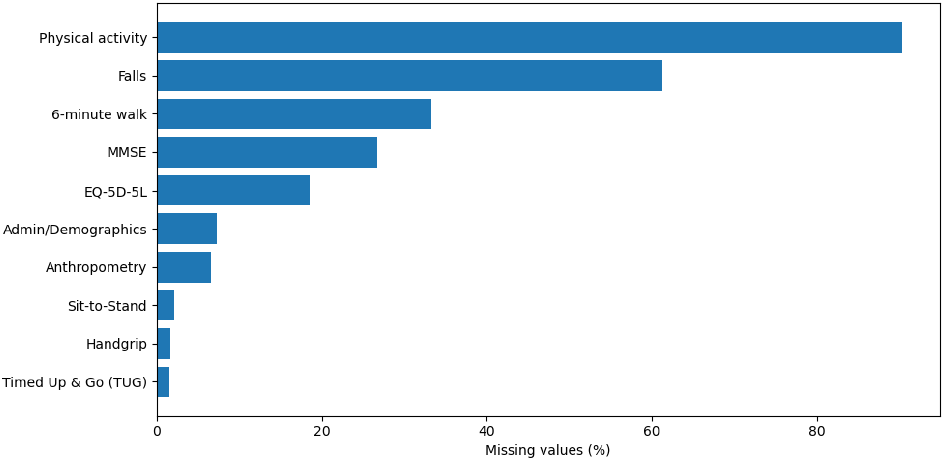
Missingness rates per section.

Missingness was then quantified per feature and grouped into bands (0–5%, 5–20%, 20–40%, and *>*40%). Features with more than 40% missingness were removed, while the remaining variables followed a treatment strategy. Given the longitudinal nature of the data, forward/backward filling within each participant was applied to selected fields to reuse information across assessment moments. Sessions with missing values in critical variables (e.g., Age or outcome related information) were removed.

Next, domain-driven deterministic rules were applied to enforce logical consistency across related variables, particularly for falls and the 6-minute walk test (e.g., ‘*no falls*’ implying zero counts and no injury/hospitalization; ‘*did not stop*’ implying zero stops and empty reasons). When a positive event was known yet the associated intensity/count was missing, medians of non-zero values were used to obtain plausible replacements. Finally, for the remaining numeric variables with low-to-moderate missingness, KNN imputation (k=10) was applied, followed by rounding/type corrections for binary, discrete, and ordinal variables, followed by a final manual validation and consistency constraints.

### 3.4 Feature Engineering

#### 3.4.1 Variable encoding and standardization

##### Encoding

To ensure compatibility with the downstream modeling pipeline, ordinal categorical variables (e.g., intensity levels and percentile bands) were numerically encoded using an order-preserving integer scale. Specific categorical fields (e.g., assessment site) were encoded via fixed integer mappings to provide a consistent representation. Finally, free-text variables were numerically encoded using neural embeddings (section 4.4.2).

##### Standardization

For all downstream models, numerical features were standardized using z-score to place variables on a comparable scale and prevent large-magnitude features from dominating distance-based procedures. Each feature *x* was transformed into a z-score, 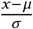, where *µ* and *σ* denote the mean and standard deviation estimated on the training data, subsequently applied to validation and test sets to avoid data leakage.

#### 3.4.2 Text representations

##### Embeddings

Free-text variables were encoded into dense numerical vectors, enabling the integration of unstructured fields into downstream analyses, and providing a principled way to quantify similarity between text responses via cosine similarity. The considered text inputs covered the fall-related descriptions, namely the reported fall circumstance, injury type, injury location, and the reason for stopping in the 6-minute walk test.

The model used to generate embeddings was the Universal Sentence Encoder, generating one fixed-length 512-dimensional vector for each entry [4]. Missing values were mapped to empty strings, and embeddings were computed efficiently by de-duplicating repeated strings.

Since the raw embeddings are high-dimensional, dimensionality reduction was subsequently tested and applied to obtain more compact representations while preserving semantic structure. Both PCA and TruncatedSVD were trained on the full embedding matrix and projected to a low-dimensional space. In order to determine the best representation, we compared the cosine similarity matrices in the original space against the reduced space, and also computed the fraction of explained variance. TruncatedSVD retained a large fraction of the variance in 10 dimensions (explained variance ≈ 0.876) and largely preserved the relative similarities between common text categories, suggesting this representation can maintain the main semantic relationships while mitigating the curse of dimensionality for subsequent modeling.

**Table 1:** Examples of LLM-assigned severity scores (0–10) for common responses.

| Text field | Text | Score |
| --- | --- | --- |
| Injury type | Bruise / hematome | 2 |
|  | Fracture | 5 |
|  | Dislocation | 6 |
| Fall context | Walking | 6 |
|  | Tripping | 4 |
|  | Dizziness / syncope-related | 10 |
| 6-min walk stop reason | Fatigue | 3 |
|  | Shortness of breath | 7 |
|  | Unable to walk | 10 |
| Injury location | Knee | 3 |
|  | Head | 8 |
|  | Skull | 10 |

##### LLM

In addition to the embedding representations, an alternative procedure based on LLM-derived score was tested to generate representations with substantially lower dimensionality. By producing a single numerical score per free-text variable, this approach avoids unnecessary dimensionality growth while enabling a more seamless integration of these features with the remaining structured features in downstream analyses, while further promoting direct interpretability.

This approach is consistent with prior work showing that large language models can act as zero-/few-shot clinical information extractors, producing structured outputs from clinical free text with minimal supervision [2].

Through this approach, the same text fields were converted into severity scores (0–10). First, the text describing the injury type was mapped to a score where minor lesions (e.g. bruises) received low values, injuries (e.g., fractures, dislocations) received intermediate values, and life-threatening events (e.g., cranial trauma, stroke, hearth attack) were assigned the highest scores. Second, the text describing the fall circumstance was scored according to a severity rationale: falls occurring during basic daily activities (e.g., walking, bathing) were rated as more severe than those during complex activities (e.g., cycling, climbing), while situations associated with acute medical symptoms (e.g., dizziness, syncope, hypotension) received the highest values. Next, the text reporting the reason for stopping the 6-minute walk test was scored by considering cardio-respiratory causes and total functional incapacity as most severe, intermediate scores for alarming pain or marked functional limitations, and lower scores for generalized discomfort. Finally, the variable indicating the injury location from a fall was mapped by assigning higher values to anatomically higher-risk sites(e.g., head, neck, spine) and lower values to peripheral locations. Unknown/empty strings were mapped to zero.

Several LLM models were tested, including Qwen models due to their open source nature. Among the tested options, the instruction-tuned model `qwen2.5:7b-instruct` was selected as it followed the prompt constraints more reliably, particularly in terms of the required output structure and score logic.

The model was fed with a comprehensive prompt describing the task, the scale meaning(0–10) and also few-shot examples covering the most common text patterns observed in the dataset to anchor consistent scoring across frequent responses.

Overall, the proposed LLM-based scoring strategy produced low-dimensional and clinically interpretable features from free-text responses, enabling their straightforward integration into the downstream analysis pipeline.

### 3.5 Associative and Trend Analysis

#### 3.5.1 Correlations

Pairwise correlation analysis is conducted to retrieve input variables exhibiting stronger monotonic or linear relationships with the outcomes, as well as to flag potential redundancy or target leakage among derived variables.

For each outcome, Spearman’s rank and Pearson’s correlations are computed between all input numeric features and targets, excluding feature-target pairs with fewer than 10 valid samples or with near-zero variance.

Table 2 highlights statistically significant, yet mostly small-to-moderate, associations between the considered outcomes and functional/anthropometric measures. For falls, the most consistent relationships involve physical function: handgrip strength shows a negative correlation (lower strength associated with higher fall occurrence), while TUG presents a positive correlation (worse performance associated with more falls); mobility and the 6-minute walk measure follow the same pattern, with poorer self-reported mobility and lower walking performance associated with higher fall occurrence. For hospital care, the associations remain directionally consistent but weaker overall. Hospitalizations exhibit the smallest effect sizes, where TUG and the 6-minute walk still appear among the top correlates, and sit-to-stand repetitions and education show negative correlations, suggesting that better mobility and higher educational attainment are linked to fewer hospitalizations. Additionally, we observed fewer falls among males, fat mass showed a positive association and sit-to-stand repetitions were negatively correlated, further reinforcing the link between reduced functional capacity and increased fall risk.

**Table 2:**
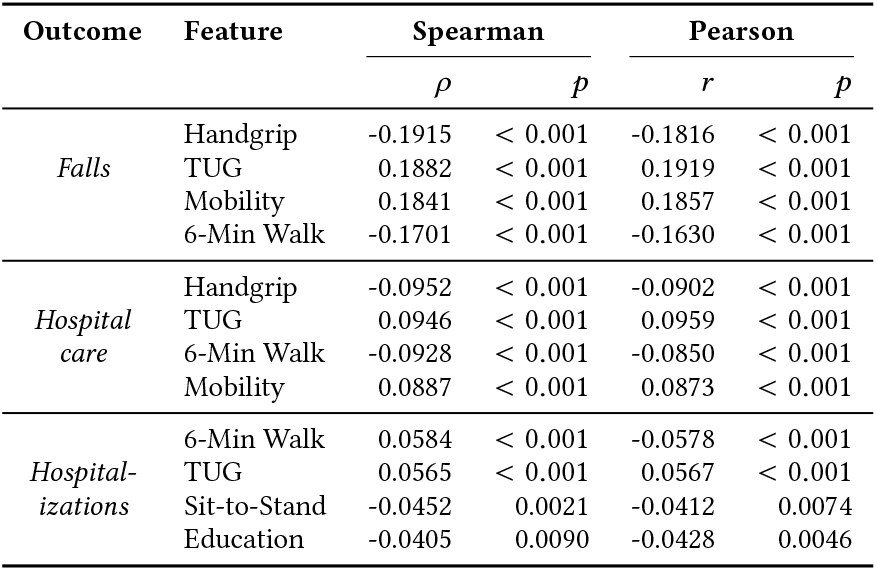
Top correlations per outcome: Spearman and Pearson.

| Outcome | Feature | Spearman |  | Pearson |  |
| --- | --- | --- | --- | --- | --- |
| | | $\rho$ | $p$ | $r$ | $p$ |
| Falls | Handgrip | -0.1915 | < 0.001 | -0.1816 | < 0.001 |
|  | TUG | 0.1882 | < 0.001 | 0.1919 | < 0.001 |
|  | Mobility | 0.1841 | < 0.001 | 0.1857 | < 0.001 |
|  | 6-Min Walk | -0.1701 | < 0.001 | -0.1630 | < 0.001 |
| Hospital care | Handgrip | -0.0952 | < 0.001 | -0.0902 | < 0.001 |
|  | TUG | 0.0946 | < 0.001 | 0.0959 | < 0.001 |
|  | 6-Min Walk | -0.0928 | < 0.001 | -0.0850 | < 0.001 |
|  | Mobility | 0.0887 | < 0.001 | 0.0873 | < 0.001 |
| Hospitalizations | 6-Min Walk | 0.0584 | < 0.001 | -0.0578 | < 0.001 |
|  | TUG | 0.0565 | < 0.001 | 0.0567 | < 0.001 |
|  | Sit-to-Stand | -0.0452 | 0.0021 | -0.0412 | 0.0074 |
|  | Education | -0.0405 | 0.0090 | -0.0428 | 0.0046 |

#### 3.5.2 Tendencies along time

Given that frailty is a dynamic clinical state, substantial insight can be gained by analysing how key clinical and functional indicators improve or deteriorate over time, rather than relying solely on cross-sectional snapshots. In particular, several widely adopted frailty definitions explicitly treat unintentional (abrupt) weight loss as a core marker[17]. In the cohort study conducted, a fraction of participants were assessed at four distinct evaluation moments(Moments 1-4) that spanned approximately two years. However, despite the availability of more than 6000 time-stamped records, only approximately 500 individuals completed assessments at all four moments, therefore, longitudinal trend analyses were restricted to this complete-case cohort.

The longitudinal analysis focused on estimating individual trends across the four evaluation moments for a wide range of clinical and functional indicators, including anthropometry, mobility tests, and muscle strength. Although signs of deterioration were observed in some variables, most notably body weight and selected functional tests the magnitude and prevalence of these changes were limited. In particular, fewer than **10%** of the participants exhibited a statistically significant decrease in body weight, and for most functional assessments, the proportion of individuals showing a significant decline is below 5%. Conversely, improvements were also detected in several of these same indicators, a pattern that is plausibly explained by the structured exercise and intervention programs in which many participants were enrolled during the follow-up period. Additionally, declines in certain measures frequently occurred in individuals who experienced a fall during the study interval, introducing ambiguity in the interpretation of causality namely, whether functional deterioration preceded or followed the fall event. Taken together the fact that only approximately 500 individuals completed four sessions, and that overall longitudinal changes were modest along two-year follow-up, these findings suggest limited statistical power to robustly outline temporal trajectories. For these reasons, and given the likelihood that the observed trends were not statistically nor clinically significant at the population level, longitudinal modeling is not further pursued in the subsequent predictive analysis.

### 3.6 Learning Setup

#### 3.6.1 Descriptive Modeling

Complementarily to the undertaken associative analysis, clustering is applied to stratify individuals according to their functional patterns. The clustering analysis is organized around a primary outcome-agnostic view and a complementary outcome-aware view. To this end, two classic methods are applied: Gaussian Mixture Model (GMM) and k-means offer complementary inductive biases: k-means provides an interpretable partition based on distances to cluster centroid representations, whereas GMM allows for probabilistic cluster membership and can capture elliptical cluster geometries and heterogeneous within-cluster variance, which are plausible in multidimensional functional assessments.

Firstly, a preprocessing step is applied, which included the removal of redundant variables and consolidating overlapping information into single composite features, in order to have a minimal yet complete feature set. Subsequently, clustering is performed under two alternative feature sets: (i) a *without-outcomes* setting, using only functional and clinical descriptors, and (ii) a *with-outcomes* setting, where fall-related outcomes are added to explicitly incorporate fall burden and perceived risk. The *without-outcomes* approach is useful to recover functional profiles in an outcome-agnostic manner, enabling downstream assessment of how outcomes differ across naturally emerging functional strata. In contrast, the *with-outcomes* approach is considered a complementary analysis whose objective is to explore stratification from an integrated view of functional characteristics and fall-related outcomes.

Given the heterogeneous nature of the dataset, feature-specific weights were introduced prior to clustering by multiplying the selected variables by knowledge-defined coefficients reflecting their intended influence in the distance/probabilistic space. This weighting scheme is used to mitigate over-segmentation driven by variables that can dominate the partition despite limited clinical interest for profiling.

Cluster quality is assessed by iterating over multiple values of *k* and selecting the partitions that achieved higher silhouette scores. To support interpretability, cluster profile maps, variable distribution plots stratified by cluster, and low-dimensional projections (e.g., PCA based embeddings) to inspect separation and overlap are undertaken.

Finally, statistical tests are performed to quantify whether clusters differed significantly across relevant variables and outcomes, complementing descriptive profiling. Statistical testing provides an objective assessment of which features are most discriminative between clusters and helps distinguish meaningful separation from noise. Appropriate tests were selected based on variable type and distribution: categorical variables are assessed via *χ*^2^ tests of independence on contingency tables, whereas continuous/ordinal variables are first screened for within-cluster normality using the Shapiro–Wilk test and then compared using parametric tests (Welch’s *t* -test for *k* = 2 or one-way ANOVA for *k >* 2) when normality held, or non-parametric alternatives (Mann–Whitney *U* for *k* = 2 or Kruskal–Wallis for *k >* 2) otherwise.

A limitation of this clustering formulation is that each record is treated as an independent observation. Therefore, participants with multiple assessment moments contribute multiple points to the clustering space, and the algorithms do not explicitly account for within-subject correlation or longitudinal dependency when forming clusters.

For completeness, additional clustering visualizations and the full set of feature-level statistical results are provided in the accompanying Supplementary Document (see Supplementary Material [37]).

#### 3.6.2 Predictive Modeling

Prediction is conducted to prognosticate multiple clinically relevant outcomes. Three classification tasks are defined: prediction of falls (*quedas*), hospital assistance (*assistência hospitalar*), and hospitalizations (*hospitalizações*).

To ensure methodological consistency and comparability across outcomes, the following classifiers are transversally applied: Extreme Gradient Boosting (XGBoost) [6], Light Gradient Boosting Machine (LightGBM) [22], CatBoost [31], Explainable Boosting Machine (EBM) [27], Support Vector Machine with RBF kernel (SVM-RBF) [10], and Multi-Layer Perceptron (MLP) [33]. These models cover a broad spectrum of learning paradigms including tree-based gradient boosting methods, kernel-based classifiers, neural networks, and inherently interpretable models. SHapley Additive exPlanations (SHAP), a unified framework based on cooperative game theory [28], is applied to acquire explanations. Explainable Boosting Machine (EBM) is excluded from SHAP analysis, as it is inherently interpretable by design. EBM is a generalized additive model that provides direct and explicit feature effect estimates, allowing model interpretation without the need for post-hoc explanations [26].

##### Handgrip Strength Regression

Handgrip strength is a validated and widely used proxy for overall muscle strength and is a key component in the assessment of physical frailty. In this study, *hand-grip strength* is modeled as a continuous variable using supervised regression techniques.

The following regression models are employed: Extreme Gradient Boosting Regressor (XGBoost) [6], Light Gradient Boosting Machine Regressor (LightGBM) [22], CatBoost Regressor [31], Explainable Boosting Machine Regressor (EBM) [27], Support Vector Regressor with RBF kernel (SVR) [10, 34], and Multi-Layer Perceptron Regressor (MLP) [33]. Model performance is evaluated using standard regression metrics, and model behavior is further analyzed using SHapley Additive exPlanations (SHAP) for all regression models, excepting Explainable Boosting Machine (EBM).

##### Sarcopenia Classification

Sarcopenia is subsequently modeled as a binary classification task using clinically established sex-specific handgrip strength thresholds. According to the revised European Working Group on Sarcopenia in Older People (EWGSOP2) consensus, low muscle strength is defined as handgrip strength below 27 kg for men and below 16 kg for women [11]. Individuals are classified as probable sarcopenic when predicted handgrip values fell below these thresholds.

For this task, a single classification model is employed in order to enable a direct and consistent comparison between the regression-based handgrip prediction and the binary sarcopenia classification, thereby avoiding redundant model evaluations.

##### Risk Aggregation via Reciprocal Rank Fusion

To integrate multiple performance criteria into a single robust risk indicator, Reciprocal Rank Fusion (RRF) score is considered for both regression and classification-based tasks. RRF is a rank-based ensemble strategy that aggregates model outputs by combining the relative ordering of predictions rather than their absolute values, thereby reducing sensitivity to scale differences and model-specific biases. For each model, individuals are ranked according to predicted risk, and a consensus score is obtained by summing the reciprocal of the adjusted ranks across models. RRF has been shown to be an effective and theoretically grounded fusion technique for combining heterogeneous ranking systems, particularly in settings involving multiple predictive models [9].

## 4 Results and Discussion

### 4.1 Frailty profiles using clustering

#### 4.1.1 Data Preparation

As clustering is performed under two settings (in the absence and presence of outcomes, cf. Section 3.6), feature selection follows a shared core, plus an outcome-dependent step. First, a set of non-clinical variables with low descriptive value, such as identifiers, assessment logistics, and background descriptors such as education level, is removed. Next, outcome related variables are handled according to the experimental setting. In the outcome-agnostic configuration, all fall-related variables are excluded to ensure that clusters reflected latent functional profiles independently of fall burden. In the outcome aware configuration, three fall-related variables are retained: the number of falls in the last 12 months, the score describing the most severe fall episode, and self-reported fear of falling, the latter being a clinically relevant marker in frailty research [12].

After this initial filtering, the feature set is further altered through feature engineering and redundancy removal. Handgrip strength, originally represented by multiple measurements, is aggregated into a derived variable, representing the best handgrip test attempt. Additionally, since two TUG tests are available and the hand-support indicator is conceptually equivalent across both variants, this information is consolidated into a single binary feature indicating whether hand support is used in either test. Additional redundancies are removed by identifying variables that conveyed essentially the same information, even when not strongly correlated, and retaining only the most interpretable measure within each redundant group. Finally, the feature set is standardized and, for pairs of variables presenting very high correlation (|*ρ*| *>* 0.9), only one variable is retained to avoid duplicate information.

Given the heterogeneous structure of the final feature set, feature-conditional weights are introduced by multiplying selected variables by predefined coefficients prior to model fitting. This weighting scheme can help prevent clusters from being dominated by variables whose influence could be disproportionate to their clinical relevance. In particular, *Gender* is down-weighted to reduce the likelihood of having partitions being primarily driven by sex-related differences. Also, the cognitive domain measured by MMSE comprises multiple sub-scores. Each MMSE component is therefore down-weighted to ensure that cognition does not receive excessive global influence merely due to being represented by many variables. Conversely, `Handgrip_Best` is up-weighted to ensure that a clinically meaningful test represented by a single feature was not diluted relative to multi-variable domains. Together, these steps yielded a standardized, de-redundant, and clinically balanced representation of functional status, which is then used as input for both clustering settings.

For completeness, additional clustering visualizations, as well as the full set of statistical results (*p*-values) are provided in the accompanying supplement (see Supplementary Material [37]).

#### 4.1.2 Outcome-Agnostic Clustering

In both clustering setups, a coarse partition with *k* = 2 is firstly analyzed to evaluate whether the cohort naturally separates into broadly robust versus frailty-related profiles. Subsequently, solutions with larger values of *k* (selected in accordance with their silhouette) are explored.

*k* = 2. The first setup with two clusters and no outcomes yields two large groups: Cluster 0 with *n* = 2906 observations and Cluster 1 with *n* = 3799. Cluster characterization is summarized in the cluster profile map (Fig. 4), which contrasts the mean standardized (z-score) values of the most discriminative features across clusters. This partition separates two functionally distinct profiles. Cluster 0 exhibits characteristics reminiscent of frailty-related deficits (e.g., lower handgrip strength, reduced 6-minute walk capacity, and poorer Timed Up and Go performance), consistent with proxies of the Fried criteria; however, given the absence of a formal frailty assessment in the dataset, this should not be interpreted as a definitive frailty diagnosis. In contrast, Cluster 1 presents systematically higher standardized values across these domains, indicating a comparatively healthier and more robust functional profile.

**Figure 4.**
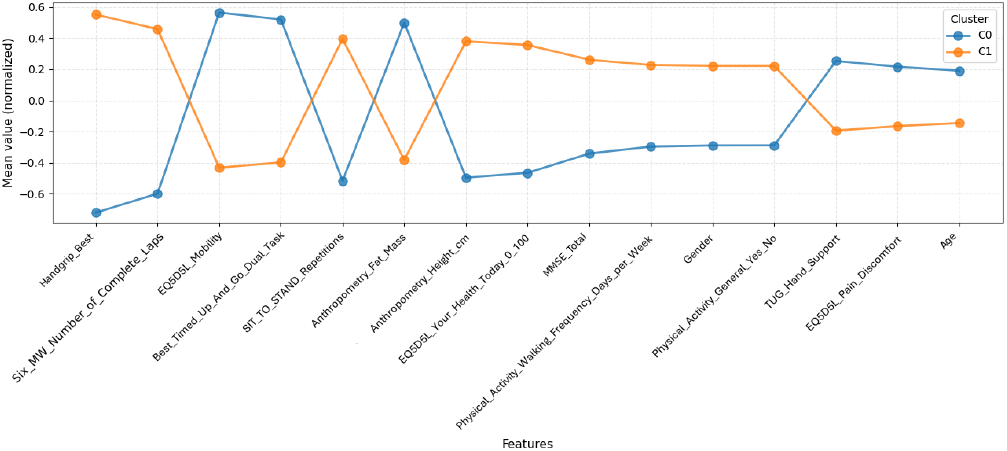
Cluster profile map for the k-means solution (*k* = 2). Mean standardized (z-score) values of the most discriminative variables across the two clusters.

Although fall-related variables are not used to form clusters in this setup, outcomes are inspected *a posteriori* to assess if the identified clusters differed in fall burden. Cluster 0 displayed a higher fall incidence, with 34% of individuals reporting at least one fall, of whom 18% sustained injuries and 9% required hospitalization. Conversely, Cluster 1 showed a lower-risk pattern with 15% reporting falls, 8.2% injuries, and 3.6% hospitalization events, supporting that outcome-agnostic clustering recovers functionally meaningful profiles that also align with differential fall burden.

Figs. 5a and 5b show the distribution of body weight and fat mass across clusters. While weight exhibits low discriminate power between functional profiles, fat mass shows a clearer separation between clusters indicating that body composition is more informative than absolute weight in capturing differences related to functional robustness.

**Figure 5.**
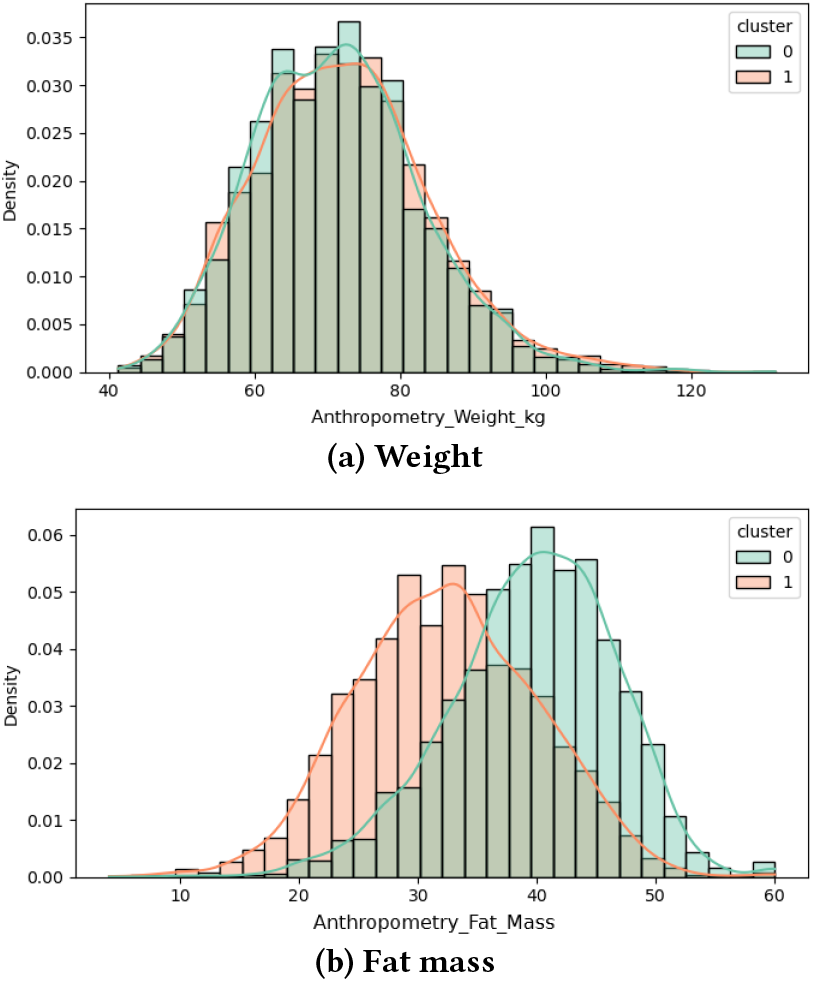
Anthropometric features across clusters (*k* = 2).

From Figs. 6a–6b, one can assess a clear shift in the distributions of sit-to-stand repetitions and handgrip strength across clusters, indicating that objective functional performance tests provide strong separation between the identified profiles.

**Figure 6.**
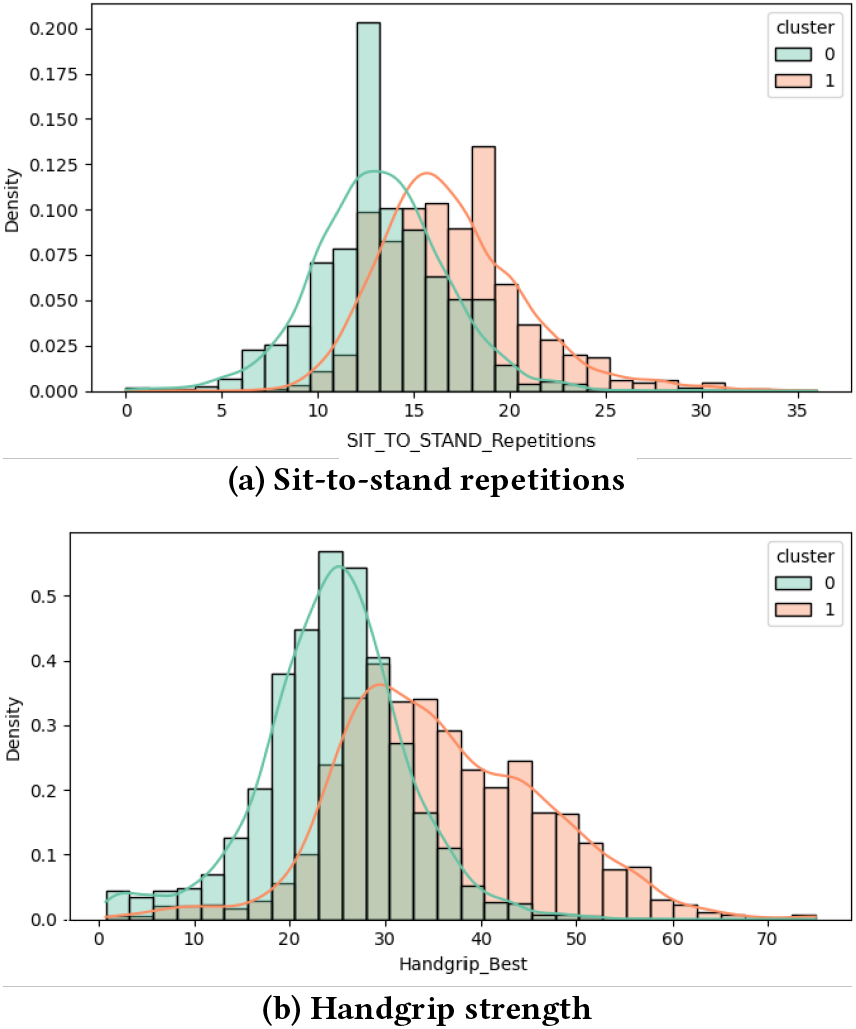
Functional performance across clusters (*k* = 2).

The statistical tests corroborate the separation observed in the profile maps and distribution plots: the most discriminative variables are related to functional performance and mobility-related domains, with extremely small *p*-values (e.g., handgrip strength, Timed Up and Go performance, sit-to-stand repetitions, and 6-minute walk capacity, all with *p <* 10^−300^), together with self-reported mobility limitations. Quality-of-life dimensions such as pain/discomfort and usual activities also show strong differences between clusters (e.g., *p* ≈ 10^−209^ and *p* ≈ 10^−165^, respectively), and global cognition remains significantly distinct (MMSE total score, *p* ≈ 10^−130^). Conversely, anthropometric measures were comparatively less informative, with visceral fat showing only modest separation (*p* ≈ 7.8 × 10^−3^) and body weight presenting the weakest separation overall (*p* ≈ 1.3 × 10^−2^), reinforcing that functional capacity is notably more discriminative than anthropometry measures.

*k* = 5. To move beyond the coarse two-group partition, a five-cluster k-means solution is adopted as the primary fine-grained outcome-agnostic representation. The solution presents a favourable silhouette score while providing clinically interpretable sub-profiles. No fall-related variable contributes to cluster formation. The resulting partition comprises Cluster 0 with 2,868 observations (42.8%), Cluster 1 with 1,863 observations (27.8%), Cluster 2 with 40 observations (0.6%), Cluster 3 with 1,844 observations (27.5%), and Cluster 4 with 90 observations (1.3%).

The subsequent analysis focuses on Clusters 1, 2, and 4, as they represent the most clinically distinctive profiles: a comparatively healthier group, a markedly vulnerable subgroup, and an intermediate functional profile, respectively. Given the comparatively small sizes of Clusters 2 and 4, their profiles should be interpreted as exploratory. Clusters 0 and 3 represent larger and less distinctive intermediate profiles and are therefore not discussed in detail.

Fig. 7 shows that the clusters are primarily differentiated by physical function and perceived health status. Cluster 1 presents the most favourable profile, characterized by better TUG performance, less reliance on hand support, higher handgrip strength, more sit-to-stand repetitions, a greater number of completed laps in the 6-minute walk test, and better self-reported quality of life, reflected by fewer EQ-5D-5L limitations and a higher VAS score. Cluster 2 exhibits the opposite pattern, combining poorer TUG performance, greater reliance on support, lower functional capacity, and worse self-rated health. Cluster 4 is particularly distinguished by walking intolerance, showing markedly higher values for stopping during the 6-minute walk test and moderately greater limitations in the EQ-5D-5L mobility and usual-activities dimensions.

**Figure 7.**
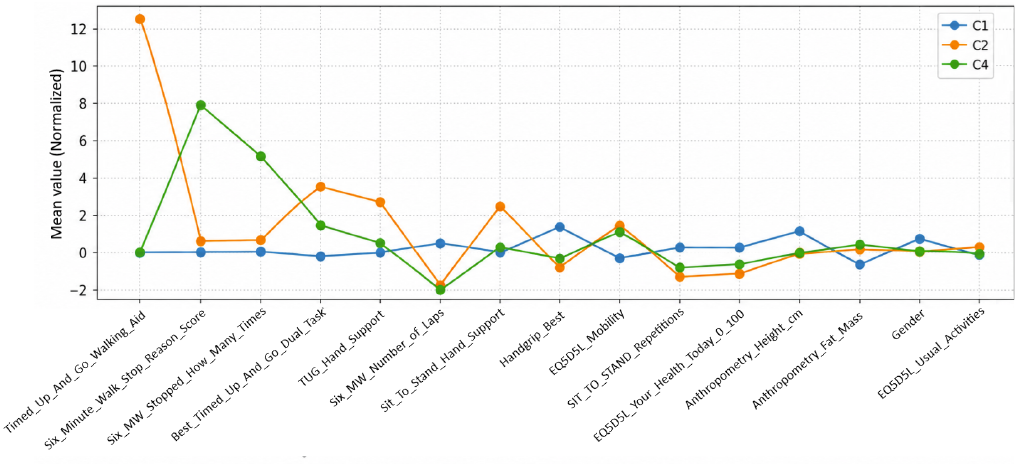
Cluster profile map (k-means, *k* = 5)

Fig. 8 shows a clear shift in dual-task TUG performance across the selected clusters. Cluster 1 presents a pronounced peak at lower completion times, indicating the most favourable mobility performance and relatively low within-cluster variability. Cluster 4 exhibits an intermediate profile, with its distribution shifted towards higher completion times and more dispersed than that of Cluster 1. Cluster 2 presents the poorest performance, characterized by a flatter and markedly right-skewed distribution extending towards substantially higher completion times, which suggests greater functional heterogeneity and a higher prevalence of mobility limitations. The statistical tests across all five clusters confirm the observed separation, with the strongest differences involving 6-minute walk stopping behaviour, dual-task TUG performance, sit-to-stand repetitions, handgrip strength, completed laps, and self-reported mobility, all with *p <* 10^−300^. Several quality-of-life dimensions also differ significantly across clusters, including self-rated health (*p* ≈ 8.5 × 10^−257^), usual activities (*p* ≈ 3.6 × 10^−256^), pain/discomfort (*p* ≈ 2.7 × 10^−234^), and self-care (*p* ≈ 1.7 × 10^−169^). Anthropometric, demographic, and cognitive variables also present significant differences, including body weight (*p* ≈ 2.0 × 10^−284^), lean mass (*p* ≈ 2.6 × 10^−121^), MMSE total score (*p* ≈ 4.7 × 10^−114^), and sex, height, and fat mass (all *p <* 10^−300^). These findings reinforce the prominent role of physical function, mobility, and perceived health status in distinguishing the identified profiles.

**Figure 8.**
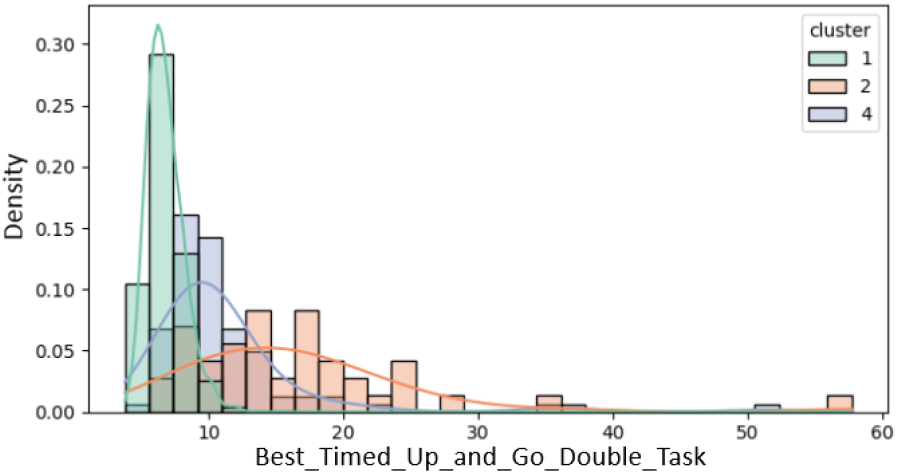
Distribution of dual-task Timed Up and Go performance across selected clusters.

#### 4.1.3 Complementary Outcome-Aware Clustering

As a complementary analysis, fall-related information—the number of falls in the last 12 months, the score associated with the most severe fall episode, and self-reported fear of falling—is incorporated into the clustering space. The objective of this setting is to explore an integrated stratification in which functional characteristics, perceived risk, and observed fall burden jointly contribute to cluster formation. Consequently, differences in fall-related variables are expected by construction and are interpreted descriptively.

*k* = 2. A GMM clustering solution with *k* = 2 clusters was selected due to its comparatively high silhouette score, indicating a well-separated partition. The resulting split was markedly imbalanced (Cluster 0: *n* = 6283; Cluster 1: *n* = 422), suggesting that outcome inclusion leads the model to isolate a relatively small and more distinctive subgroup rather than producing two broad, similarly sized profiles.

Fig. 9 shows a clear separation between a large reference cluster and a smaller integrated higher-vulnerability profile. Cluster 1 combines poorer TUG performance, greater reliance on hand support, reduced 6-minute walk capacity, and greater limitations in EQ-5D-5L mobility, usual activities, and self-care. Since fall-related variables also contribute directly to cluster formation, this cluster additionally presents a higher number of falls, greater fall severity, and a higher prevalence of fear of falling. Overall, this complementary analysis illustrates how functional impairment and fall-related burden can be jointly integrated into a single stratification. The resulting two-cluster partition is retained for the subsequent longitudinal transition analysis.

**Figure 9.**
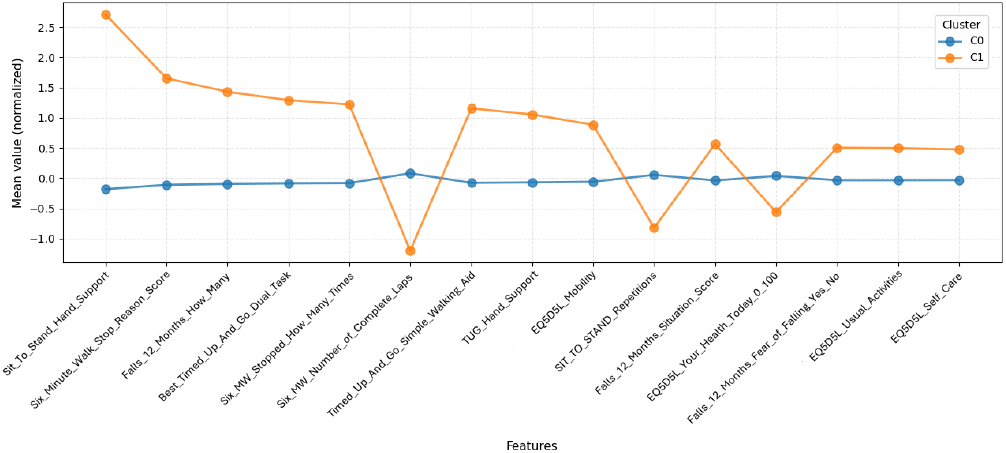
Cluster profile map (GMM, *k* = 2).

With the variable distributions across clusters, it is possible to visually examine which features differentiate average profiles from high risk profiles, in the given population.

Together, Figs. 10a and 10b indicate that the 6-minute walk test provides a two-way separation between clusters: the total number of completed laps shows a clear shift, with the more vulnerable cluster achieving fewer laps on average, while stopping behavior is also highly discriminative, as the robust cluster presents almost exclusively zero stops (close to 100%).

**Figure 10.**
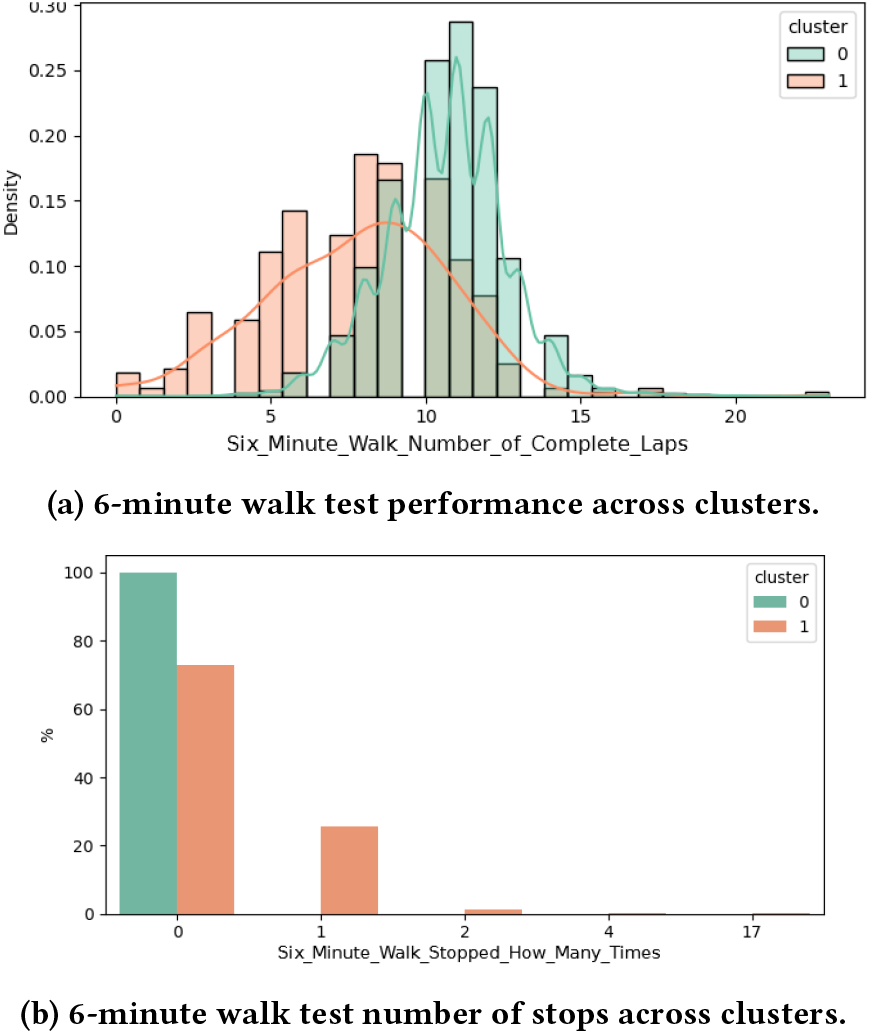
6-minute walk test outcomes across clusters (*k* = 2).

The cluster drivers are further subjected to statistical testing. The 6-minute walk stopping behavior is among the strongest discriminators (stopping motive and number of stops, both with *p <* 10^−300^), together with indicators of hand support on TUG tests (*p* ≈ 1.2 × 10^−286^) and reduced walking capacity (completed laps, *p* ≈ 6.2 × 10^−89^). Outcome variables also significantly differentiate clusters, including the number of falls in the last 12 months (*p* ≈ 2.6 × 10^−41^), the worst-fall severity score, besides being a hand-crafted feature (*p* ≈ 3.6 × 10^−32^), and fear of falling (*p* ≈ 8.0 × 10^−27^), indicating that the isolated subgroup concentrates substantially higher fall burden and perceived risk. Cognitive status remains statistically distinct but less dominant (MMSE total, *p* ≈ 1.1 × 10^−13^), while anthropometry variables show once again little discrimination power (fat mass, *p* ≈ 8.9 × 10^−6^; body weight, *p* ≈ 8.3 × 10^−2^).

#### 4.1.4 Cluster Transitions and Longitudinal Analysis

To complement the cross-sectional clustering results, cluster transitions are further examined from a longitudinal stance. Since the dataset contains repeated observations for the same individuals across up to four assessment moments, cluster membership can be tracked over time to quantify switching between functional profiles.

The analysis focuses on the outcome aware GMM solution with *k* = 2, which yields an interpretable stratification composed of a large reference cluster representing the cohort average, and a smaller cluster capturing a more vulnerable, higher fall incidence subgroup.

After restricting the analysis to individuals with at least two assessment moments (*n* = 2050), it was noted that cluster membership is largely stable over time: 86.4% of individuals remained consistently in the healthier cluster and 1.4% remained consistently in the more vulnerable cluster. Overall, 12.2% experienced at least one transition between clusters, typically a single switch, with transitions occurring in both directions at comparable rates (52.2% from healthy to vulnerable and 47.8% from vulnerable to healthy).

To contextualize cluster transitions in relation to the annual physical-intervention programme, we stratified changes in cluster membership by assessment pair. Across consecutive assessments, membership remained highly stable in the robust profile (Robust→Robust: 89.0% in 1 → 2, 86.7% in 2 → 3, and 89.1% in 3 → 4). Deteriorations from robust to vulnerable stayed uncommon (Robust → Vulnerable: 6.1% in 1 → 2, 4.8% in 2 → 3, and 4.1% in 3 → 4), while improvements from vulnerable to robust occurred at comparable magnitudes (Vulnerable→Robust: 3.0% in 1→2, 6.1% in 2 → 3, and 4.8% in 3 → 4). Persistent vulnerability remained rare across pairs (Vulnerable→Vulnerable: 1.9–2.4%). Similar patterns also appear in non-consecutive comparisons (1 → 3, 1 → 4, 2 → 4), suggesting that transitions do not concentrate in a single time window; instead, a small subset of participants shows meaningful profile changes over time, which may reflect variability in adherence and/or intercurrent events rather than programme timing alone.

To investigate which within-subject changes are associated with transitions from the healthier to the more vulnerable cluster, standardized feature deltas were computed between consecutive assessment moments,

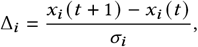

where *x*_*i*_ *t* and *x*_*i*_ *t* 1 denote two consecutive measurements for the same individual, and *σ*_*i*_ is the standard deviation of variable computed over the full dataset.

For transitions from Healthy to High Risk (Appendix, Table A1), the largest standardized increases are dominated by increased reliance on hand support during sit-to-stand and TUG, stopping during the 6-minute walk test, and a concurrent increase in fall frequency. In parallel, the largest decreases occur in endurance and strength-related measures, with reductions in 6-minute walk distance/laps and handgrip strength, suggesting that deterioration is primarily expressed through worsening mobility capacity, greater need for assistance, and increased fall burden.

Conversely, transitions from High Risk to Healthy (Appendix, Table A2) are characterized by improvements in walking capacity and functional performance, as reflected by increases in completed laps in the 6-minute walk, better TUG performance, and small gains in lean mass and self-rated health, together with a higher prevalence of general physical activity. The strongest decreases mirror the Healthy-to-High Risk pattern, showing reduced reliance on hand support, fewer 6-minute walk stops (and lower stopping-reason scores), reduced use of walking aids, and a decrease in the number of falls, indicating that recovery is associated with improved endurance, reduced functional dependence.

From a clinical perspective, these patterns provide actionable signals: a patient showing a new need for hand support during sit to stand/TUG and the onset of stopping behavior in the 6-minute walk constitute strong determinants of functional deterioration, whereas improved walking capacity (more completed laps and better TUG performance), the surge of lean mass, fewer 6-minute-walk stops, and a reduction in fall frequency can be interpreted as practical “green flags”.

Overall, the primary outcome-agnostic clustering analysis separated coherent functional profiles and highlighted physical performance, mobility, strength, and perceived health as the main dimensions of heterogeneity. These profiles also aligned *a posteriori* with different fall-related patterns. The complementary outcome-aware analysis provided an integrated stratification in which functional descriptors, observed fall burden, and perceived risk jointly contributed to cluster formation.

Across analyses, the most consistently discriminative variables encompass objective mobility and strength indicators, namely TUG-based performance and reliance on support, sit-to-stand performance and need for hand support, and 6-minute walk capacity and stopping behaviour, together with self-reported mobility limitations and broader EQ-5D-5L health-status dimensions. Fall-specific variables (number of falls, severity of the worst fall episode, and fear of falling) are, when included, among the strongest separators, reflecting their direct contribution to the integrated outcome-aware partition. In contrast, anthropometric measures such as body weight tend to show weaker discrimination between groups.

A longitudinal transition analysis further reveals within-subject movements between profiles; nevertheless, given the limited number of time points per individual (up to four), the longitudinal component should be primarly interpreted as a constrained additional insight, complementary to the ground cross-sectional stratification.

### 4.2 Predictive Modeling

#### 4.2.1 Data preparation

As some functional tests are repeated per visit, we standardized these replicates by keeping a single clinically meaningful summary per test: the best performance (maximum) across attempts, which aligns with common clinical practice for functional testing and improves comparability across participants [39]. The target predictors do not rely on long-term trajectory modeling given the primary focus on between-session prognostics. In this context, and given a specific outcome, the earliest available session-pair for individuals without the outcome is selected and, for cases, the first session-pair at which the outcome becomes positive. This ensures that predictors reflect information available up to the earliest positive event while also helping mitigate class imbalance. Finally, we applied a stratified hold-out split (80/20) to preserve class balance between training and test sets.

#### 4.2.2 Prediction of Fraility-related Risks

Table 3 shows moderate predictive performance for fall occurrence, with AUROC values ranging from 0.636 to 0.696. MLP achieves the highest AUROC (0.696), PR-AUC (0.315), and F1-score (0.423), whereas EBM achieves the highest recall (0.911). It is worth noting that the decision threshold is not fixed at 0.5 instead, it’s selected to minimize a misclassification cost (prioritizing sensitivity by reducing false negatives). As these differences are small and could be driven by the particular train/test split, hold-out observatios are complemented by a stratified 10-fold cross-validation and fold-wise statistical testing. Using paired tests on the per-fold AUCs, we found that EBM is consistently the strongest predictor and is statistically superior to the remaining models (with statistically significant p-values), with CatBoost also ranking among the best and is superior to the other alternatives. Despite these results, the confusion matrices indicate a non-negligible number of false negatives, which is expected because falls are intrinsically uncertain events and can be triggered by unobserved, context-dependent factors (e.g., environment and acute incidents), limiting the maximum achievable sensitivity from baseline assessments alone.

**Table 3:** Falls - average performance comparison across models, given an adjusted threshold.

| Model | AUROC | PR-AUC | F1-score | Recall |
| --- | --- | --- | --- | --- |
| Logistic Regression | 0.636 | 0.260 | 0.383 | 0.810 |
| CatBoost | 0.674 | 0.308 | 0.395 | 0.582 |
| XGBoost | 0.658 | 0.284 | 0.369 | 0.608 |
| EBM | 0.649 | 0.259 | 0.376 | **0.911** |
| SVM (RBF) | 0.668 | 0.280 | 0.403 | 0.595 |
| MLP | **0.696** | **0.315** | **0.423** | 0.709 |

Figures 11 and 12 show that both models converge on clinically plausible drivers of fall risk that map onto core frailty-related domains, including gender (there is higher frailty burden in women), handgrip strength (`HANDGRIP_BEST`), and self-reported function via EQ-5D-5L mobility (`EQ5D5L_MOBILIDADE`). In the CatBoost SHAP summary plot each point corresponds to one participant, the horizontal axis reports the SHAP value (the signed contribution of the feature to the model output for that individual), and the color encodes the feature value (blue = low, red = high), allowing simultaneous inspection of directionality and inter-individual variability. In addition to these established descriptors, the models also highlight cognitive/visuoconstructive performance (copy of a drawing; `MMSE_COPIA_DESENHO_1`), which is noteworthy given that this task is not typically regarded as a primary frailty marker. CatBoost further emphasizes performance-based functional tests, including endurance and mobility proxies such as the Six-Minute Walk Test(e.g., distance and number of laps) and TUG variants (simple and dual-task). By contrast, the EBM importance profile exhibits a distinctive behavior: beyond main effects, EBM explicitly models and reports pairwise interactions, which yields prominent combined terms (e.g., `ANTROPOMETRIA_Gordura_visceral` & HANDGRIP_BEST) and a comparatively stronger focus on anthropometric indicators (fat mass, visceral fat, lean mass, stature) rather than purely functional tests. Overall, the two approaches capture complementary facets of frailty-related vulnerability, with CatBoost prioritizing functional performance (TUG, Six-Minute Walk Test) and EBM highlighting body-composition measurements and their interactions with strength.

**Figure 11.**
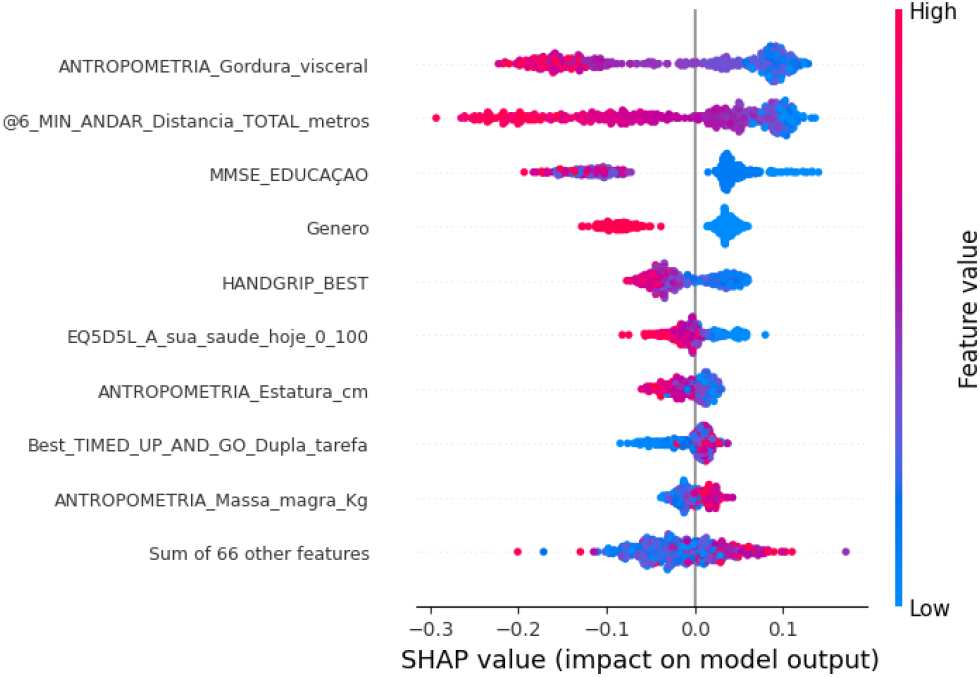
SHAP summary plot of the CatBoost model showing the top-10 features for fall risk prediction. Color indicates the feature value (blue = low, red = high).

**Figure 12.**
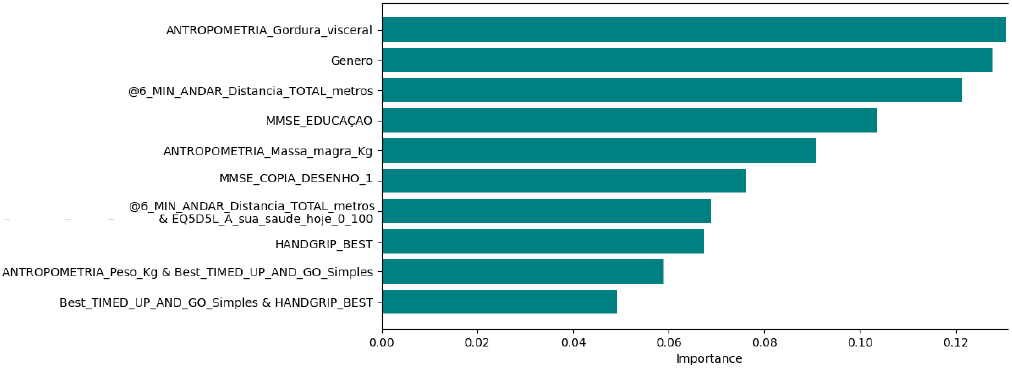
Global feature importance of the Explainable Boosting Machine (EBM) for fall risk prediction (top-10).

As highlighted in Tables 4 and 5, *assistência hospitalar* and *hospitalizações* are more difficult to prognosticate than falls, largely because these outcomes are more imbalanced in the dataset and can be strongly influenced by additional externalities (e.g., the severity of an incident or injury, an individual’s decision to seek hospital care, and contextual/social factors such as home safety or the availability of informal support at home). While AUROC values remain moderately positive, the relatively low F1-scores indicate that models often need to trade precision for sensitivity, producing a number of false positives to correctly classify a meaningful fraction of positive cases. In terms of explainability, the top-10 features are broadly similar with the fall-prediction model with moderate changes in ordering, suggesting that the models are primarily capturing the dependency between falls and medical assistance and not the desired risk features. In line with this, pairwise statistical comparisons to identify a clearly superior model were largely inconclusive, with few consistent significant differences across folds, reinforcing that these endpoints are not being captured with robust, discriminative patterns in the current feature set. Consequently, these results are best interpreted as exploratory in the context of the current dataset. The consensus risk score is constructed as a democratic aggregation of the individual classifiers, aiming to capture the extent to which different models *agree* that a participant is at higher risk. Concretely, each model produces a risk estimate for the same individual, and these outputs are first mapped onto a common scale. The consensus score then combines them into a single value (0–100%) that increases when multiple models simultaneously assign higher risk and decreases when the models assign low risk or disagree. As the underlying models can differ in calibration and confidence, this consensus score should not be interpreted as a simple arithmetic mean of probabilities, but rather as an agreement-weighted summary of model evidence.

**Table 4:** Hospital assistance - average performance comparison across models, given an adjusted threshold.

| Model | AUROC | PR-AUC | F1-score | Recall |
| --- | --- | --- | --- | --- |
| Logistic Regression | 0.632 | 0.099 | 0.178 | 0.138 |
| XGBoost | 0.683 | 0.106 | 0.218 | 0.379 |
| CatBoost | **0.689** | 0.138 | 0.195 | **0.655** |
| EBM | 0.634 | 0.106 | 0.200 | 0.207 |
| SVM (RBF) | 0.639 | **0.159** | **0.231** | 0.207 |
| MLP | 0.663 | 0.126 | 0.199 | 0.552 |

**Table 5:** Hospitalizations - average performance comparison across models, given an adjusted threshold.

| Model | AUROC | PR-AUC | F1-score | Recall |
| --- | --- | --- | --- | --- |
| Logistic Regression | 0.659 | 0.032 | 0.095 | 0.200 |
| XGBoost | 0.696 | 0.019 | 0.046 | 0.400 |
| CatBoost | **0.734** | 0.048 | 0.154 | 0.400 |
| EBM | 0.701 | 0.042 | 0.160 | 0.400 |
| SVM (RBF) | 0.498 | 0.010 | 0.026 | **0.800** |
| MLP | 0.641 | **0.049** | **0.174** | 0.400 |

The density curves in Figure 13 show substantial overlap between fallers and non-fallers around the main peak, indicating that a large fraction of participants receive similar consensus scores regardless of the observed outcome. This overlap is consistent with falls being influenced by context-dependent and partly unobserved factors. Moreover, aggregating models can introduce additional uncertainty since disagreement across models broadens the score distribution and can blur separation, since the consensus combines shared signal but also shared noise and calibration differences.

**Figure 13.**
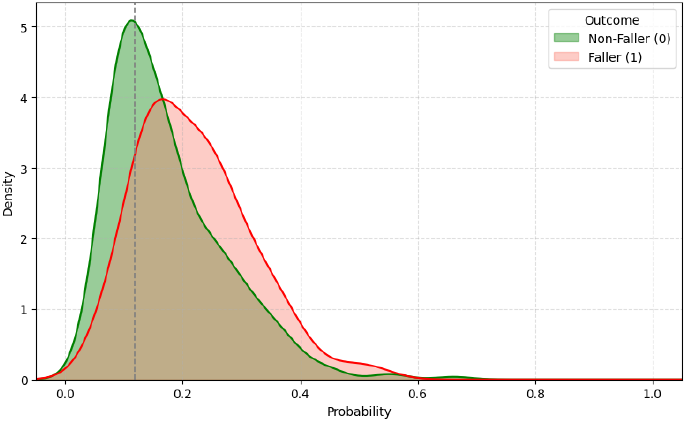
Density distribution of the consensus risk score for fallers *vs*. non-fallers.

Figure 14 provides as analogous view using the model with the best results (EBM), where the horizontal axis corresponds directly to a predicted probability. Although a pronounced overlap remains in the central region, indicating that many individuals fall into an ambiguous risk zone the EBM distribution shows a clearer separation than the consensus score, with fallers exhibiting a more evident shift toward higher predicted probabilities. This suggests that, in this setting, the consensus aggregation smooths and dilutes part of the discriminative signal, whereas the single-model EBM retains a sharper risk gradient.

**Figure 14.**
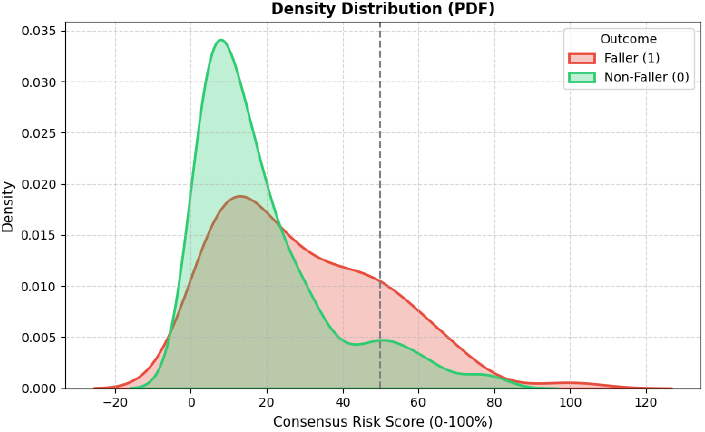
Density distribution of EBM predicted probabilities for fallers vs. non-fallers, with the best threshold.

#### 4.2.3 Handgrip Regressor and Sarcopenia Classification

Figure 15 and table 6 illustrates the performance of the CatBoost regressor, which achieved the best overall results for handgrip prediction. Beyond having the lowest average MAE across folds, pairwise statistical comparisons of MAE indicate that CatBoost significantly outperforms key alternatives (e.g., XGBoost, LightGBM, SVR, and also EBM), supporting its selection as the strongest regressor under our evaluation protocol. Nevertheless, the fit remains moderate (e.g. *R*^2^ ≈ 0.49) and dispersion around the diagonal is expected, since estimating an individual’s muscle strength from heterogeneous clinical and functional variables rather than direct physiological measurements of force production inevitably introduces uncertainty and residual variability.

**Table 6:** Handgrip regression: model performance comparison.

| Model | MAE (kg) | RMSE (kg) |
| --- | --- | --- |
| Linear Regression | 3.73 | 5.33 |
| XGBoost Regressor | 3.58 | **5.23** |
| CatBoost Regressor | **3.57** | 5.23 |
| EBM Regressor | 3.59 | 5.28 |
| SVR (RBF) | 3.76 | 5.39 |
| MLP Regressor | 4.09 | 5.67 |

**Figure 15.**
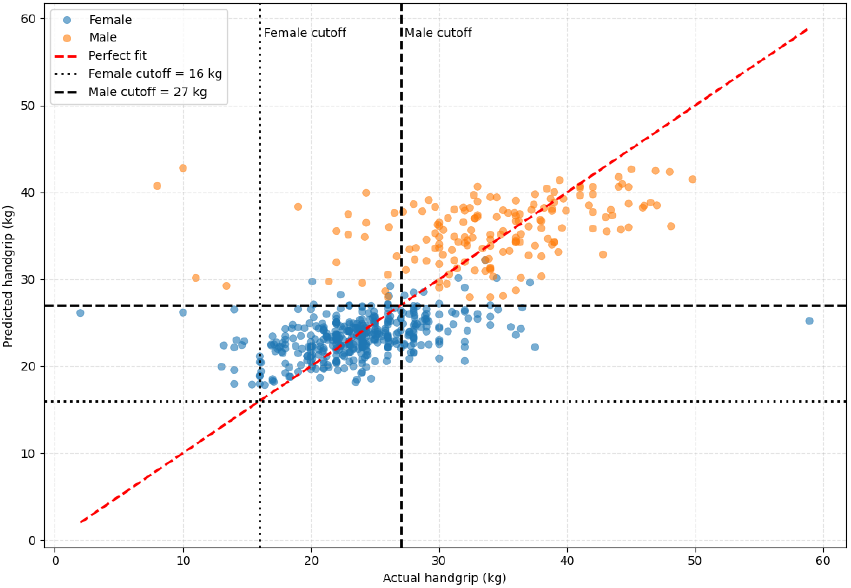
CatBoost regressor: actual vs. predicted handgrip strength by sex with sarcopenia cuttofs.

To connect the regression outputs to a clinically meaningful end-point, Fig. 15 also overlays sex-specific sarcopenia cutoffs as vertical and horizontal reference lines (female: 16 kg; male: 27 kg). The vertical lines indicate the clinical threshold applied to the *true* handgrip value (x-axis), whereas the horizontal lines apply the same threshold to the *predicted* handgrip value (y-axis), so that the bottom-left quadrant (below the corresponding cutoff on both axes) represents cases that would be classified as probable sarcopenia both clinically and by the model. In practice, the plot shows that predicted values concentrate around the central range of grip strength for each sex, without any predictions falling below the cutoffs, which means that sarcopenia cases are not identified when thresholding the regressor output. This behavior is consistent with a regression to the mean effect in an imbalanced setting, since low-strength observations are relatively scarce and require a large deviation from typical values, the model is penalized for extreme predictions and tends to remain close to the conditional average.

To address the poor performance of deriving probable sarcopenia labels by thresholding the handgrip regressor outputs, we trained a dedicated CatBoost classifier to directly predict sarcopenia as a binary outcome. We selected the operating point by tuning the decision threshold for higher sensitivity to class 1 resulting in a best threshold of 0.406, an AUROC of 0.798, and a recall for sarcopenia of 0.792 (38/48). Despite the clear improvement in detecting positive cases, precision remains low (0.195) and the F1-score is 0.313, indicating that the model still produces a substantial number of false positives, a trade-off that is expected when prioritizing sensitivity in an imbalanced screening setting. The confusion matrix in Fig. 16 shows that the classifier correctly identifies most sarcopenia cases (TP=38) while missing relatively few (FN=10), at the cost of more healthy individuals being flagged (FP=157). In practice, this behavior can still be highly useful as a *screening* tool. By triaging who should receive confirmatory assessment, this behaviour can reduce the number of full diagnostic tests required and help identify higher-risk individuals in resource-limited or remote settings using only a small set of simple measurements (e.g., basic functional tests and anthropometrics).

**Figure 16.**
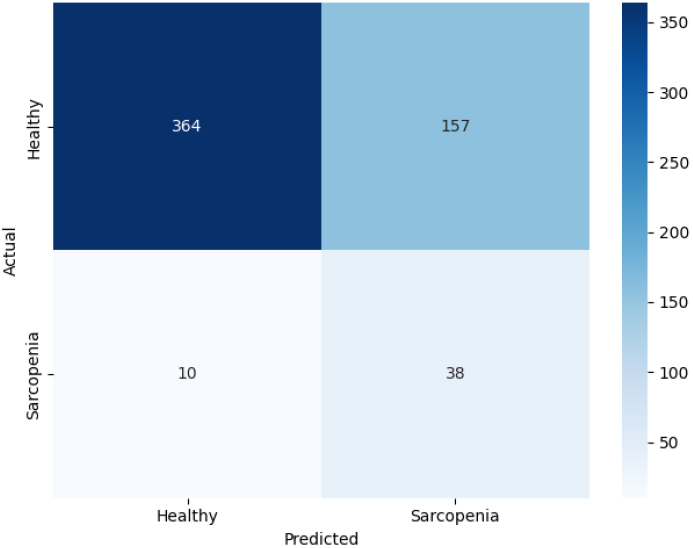
Confusion matrix for the CatBoost sarcopenia classifier at the threshold tuned for higher Recall(1) (F2-optimal).

Figure 17 shows the CatBoost SHAP summary for the sarcopenia classifier (top 10 features). The plot highlights that several performance-based functional tests carry strong predictive signal for handgrip strength, notably sit-to-stand repetitions and TUG variants, alongside mobility/endurance proxies such as Six-Minute Walk Test distance. Together with anthropometric variables (e.g., stature, weight, lean mass) and age, these results reinforce that the physical assessments collected in the cohort are indicators of overall strength capacity and can be leveraged to estimate handgrip even when direct dynamometry is not used as an input.

**Figure 17.**
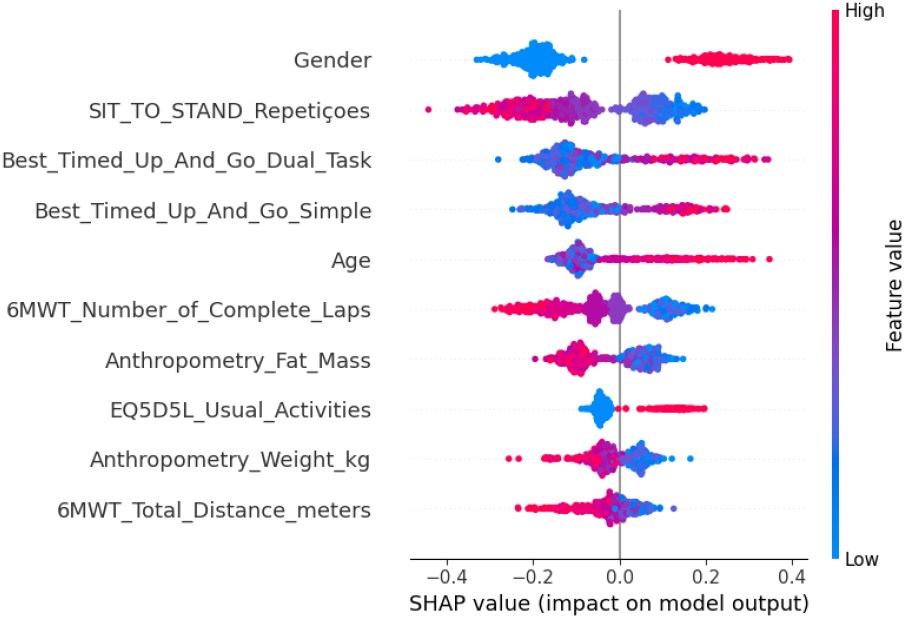
SHAP summary plot of the CatBoost sarcopenia classifier showing the top 10 most influential features. Color indicates the feature value (blue = low, red = high).

## 5 Conclusion

This work applied explainable machine learning to characterize frailty-related vulnerability and to prognosticate fall- and sarcopenia-related outcomes in older adults using data from a large-scale physical-activity follow-up program. By combining unsupervised and supervised approaches, we obtained consistent and clinically interpretable insights. Clustering revealed coherent functional profiles: when outcomes were excluded, groups were mainly separated by objective physical performance and self-reported function, and these profiles aligned a posteriori with different fall burdens. Clustering revealed coherent functional profiles: the primary outcome-agnostic analysis separated groups mainly according to objective physical performance and self-reported function, with these profiles aligning *a posteriori* with different fall burdens. A complementary outcome-aware analysis further illustrated how functional limitations, observed fall burden, and fear of falling can be jointly integrated into a descriptive stratification.

Supervised models achieved moderate discrimination for fall occurrence (AUROC = 0.636–0.696), and explainability analyses consistently highlighted domains related to strength, mobility/function, and endurance. Interpretable boosting models added value by exposing meaningful interactions between body composition and strength. In contrast, hospital assistance and hospitalization were substantially harder to predict and did not show robust, discriminative patterns in the available features, supporting their interpretation as exploratory endpoints in this dataset.

Handgrip strength is estimated with moderate accuracy, but deriving sarcopenia labels by thresholding regression outputs proved ineffective due to regression-to-the-mean effects. A dedicated sarcopenia classifier improved case detection (high recall) at the cost of many false positives, consistent with a screening-oriented setting where sensitivity is prioritized over precision. Overall, these findings suggest that explainable models built on routine, low-cost assessments can support community and primary care workflows by providing interpretable risk stratification and helping prioritize individuals for targeted follow-up.

Since the municipal initiative is still ongoing, the current follow-up horizon is limited, and participant retention across assessment waves is incomplete, which constrains the depth of longitudinal analyses. A second limitation is the substantial missingness observed across multiple variable groups, while extensive cleaning and imputation were necessary to build consistent modeling tables, the reliance on the treatment pipeline can reduce statistical robustness and attenuate true associations. Future data collection would therefore benefit from increased rigor and standardization in data entry and assessment procedures, particularly for manually entered and questionnaire-based fields, to reduce missingness and improve reliability. Third, the cohort is drawn from a municipal physical activity program and is therefore predominantly active, introducing selection bias and limiting generalizability to frailer, institutionalized, or less active populations. Broadening recruitment or validating externally on less active individuals would strengthen the applicability of the drawn putative conclusions. Finally, the absence of an explicit clinical frailty diagnosis prevents direct validation against established frailty labels, increasing uncertainty when interpreting frailty-related clustering and predictions.

Future work will incorporate standardized frailty instruments or clinician-rated annotations to enable direct benchmarking against established clinical definitions. Beyond these dataset-level extensions, an important translational direction is to strengthen sarcopenia-related modeling by combining handgrip strength with simple anthropometric circumferences (e.g., mid-upper arm, calf, and thigh circumferences). Together with a small set of targeted personal and functional questions, these low-burden measures could support the development of lightweight at-home screening tools to identify high-risk individuals early, facilitate timely referral for confirmatory assessment, guide rehabilitation protocols, and ultimately provide practical decision support and resource management for community and primary care settings.

## Data Availability

This study used only openly available datasets that were released to the public prior to the initiation of the study. Specifically, we used BRISC2025 (brain tumor MRI, https://arxiv.org/abs/2501.01836), ChestX-ray8 (https://nihcc.app.box.com/v/ChestXray-NIHCC), and MVTec Anomaly Detection (https://www.mvtec.com/company/research/datasets/mvtec-ad), all of which are publicly accessible for research purposes. No new human data were collected and no restricted or application-based datasets were used.

## Funding

This work was supported by FCT - Fundação para a Ciência e Tecnologia in the scope of the project UID/04378/2025 (DOI identifier 10.54499/UID/04378/ 2025), and UID/PRR/04378/2025 (DOI identifier 10.54499/UID/PRR/04378/ 2025), of the Research Unit on Applied Molecular Biosciences - UCIBIO and the project LA/P/0140/2020 (DOI identifier 10.54499/LA/P/0140/2020) of the Associate Laboratory Institute for Health and Bioeconomy - i4HB, INESC-ID plurianual UID/50021/2025 (DOI identifier 10.54499/UID/50021/2025); and the project FRAIL (2024.07266.IACDC). CIAFEL and ITR funded by FCT (CIAFEL: UIDB/00617/2020 and ITR: LA/P/0064/2020).

## Ethics declarations

Ethical approval was obtained from the Municipality of Vila Nova de Famalicão for the analysis of the anonymized dataset.

## Competing interests

The authors declare that they have no competing interests.

## Acknowledgments

The authors thank Famalicão’s municipal program *“Mais e Melhores Anos”* for the anonymized data provision and support.

## Appendix

**Figure A1.**
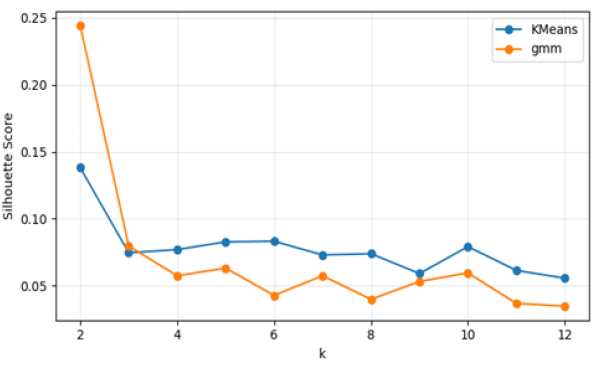
Silhouette Score for varying *k* values

**Table A1:** Largest standardized within-subject changes for transitions from Healthy to Frail.

| Feature (short description) | $\Delta_i$ |
| --- | --- |
| *Top 5 increases* |  |
| Hand support in sit-to-stand | 3.23 |
| Number of falls (last 12 months) | 1.21 |
| Stopped during 6-min walk | 1.20 |
| 6-min walk stopping reason score | 1.10 |
| Hand support in TUG (simple) | 0.94 |
| *Top 5 decreases* |  |
| 6-min walk total distance (m) | -0.48 |
| 6-min walk completed laps | -0.42 |
| Handgrip strength (best) | -0.24 |
| Lean mass (kg) | -0.21 |
| MMSE: figure copying | -0.12 |

**Table A2:** Largest standardized within-subject changes for transitions from Frail to Healthy.

| Feature (short description) | $\Delta_i$ |
| --- | --- |
| *Top 5 increases* |  |
| 6-min walk completed laps | 0.27 |
| TUG (simple) percentile | 0.20 |
| Lean mass (kg) | 0.13 |
| General physical activity (yes/no) | 0.05 |
| Self-rated health (EQ-5D VAS, 0–100) | 0.04 |
| *Top 5 decreases* |  |
| Hand support in sit-to-stand | -3.32 |
| Stopped during 6-min walk | -1.50 |
| 6-min walk stopping reason score | -1.18 |
| Walking aid in TUG (simple) | -0.95 |
| Number of falls (last 12 months) | -0.86 |

## Notes

### Competing Interest Statement

The authors have declared no competing interest.

### Author Declarations

Ethics commitee/IRB of Municipality of Vila Nova de Famalicao, Program "Mais e Melhores Anos", gave ethical approval for this work.

### Summary of Updates

Title and some additional text updated.

